# Vaccine efficacy against naturally asymptomatic infections: A novel estimand for quantifying vaccine effects

**DOI:** 10.1101/2025.08.20.25334070

**Authors:** Elizabeth Rogawski McQuade, Razieh Nabi, Allison Codi, Natalie Dean, Marc Lipsitch, David Benkeser

## Abstract

The naive approach to estimating the effects of a vaccine on asymptomatic infections, which compares the risk of asymptomatic infection among vaccinated and unvaccinated individuals, can be misleading because it is comprised of two effects: the vaccine preventing asymptomatic infections and the vaccine converting symptomatic to asymptomatic infections. When the latter effect is strong, vaccines can appear harmful with respect to asymptomatic infections. Using a causal principal stratification framework, we formalize an estimand, vaccine efficacy against naturally asymptomatic infection (VE_NAI_), that describes the effectiveness of a vaccine in preventing asymptomatic infections among individuals who would naturally (i.e., in the absence of vaccine) be expected to be asymptomatic. This estimand excludes vaccine effects that convert symptomatic cases to asymptomatic infections, and we demonstrate how this makes it a more natural analogue of the usual vaccine efficacy estimands against infection and symptomatic disease. We describe the assumptions under which this estimand can be identified and estimated from randomized and observational studies. We further identify and estimate bounds that do not require cross-world independence assumptions and characterize sensitivity analyses around the main assumption needed for identification. Finally, we apply these methods to a randomized trial of the COVID-19 mRNA-1273 vaccine. In this trial, VE_NAI_ was higher than standard estimates of efficacy against asymptomatic infections and was similar in magnitude to efficacy against any infection. Reporting VE_NAI_ in vaccine trials in addition to other vaccine effects would improve interpretability, could broaden understanding of vaccine impact on transmission, and provide insights into immunological mechanisms.

## 1 Introduction

Many vaccines effectively prevent illness and/or severe disease but may not provide protection against infection, also known as sterilizing immunity.^1^ This is especially true for respiratory infections, including influenza and COVID-19. While prevention of illness is typically the main public health goal, a vaccine that prevents infection has the major benefit of reducing onward spread.^2,3^ For vaccines with low vaccine efficacy (VE) against infection, additional non-pharmaceutical interventions may be necessary to mitigate transmission.

For individuals who may experience only mild or asymptomatic infections, a relevant question is whether and how they would benefit from vaccination. If infected, these individuals may still contribute to onward transmission, potentially even more so than symptomatic cases since asymptomatic individuals are unlikely to know they are infected and therefore less likely to take precautionary steps such as self-isolation. Thus, a vaccine that reduces the risk of asymptomatic infections may prevent community spread,^4,5^ and this benefit may be a key motivator to vaccinate individuals who are at low risk of symptomatic infection or severe disease. Furthermore, epidemiologic evidence that a vaccine prevents infections may provide insight into immunology that could inform future vaccine development and optimization.^6^ Such evidence could indicate, for example, that the vaccine induces relevant elements of mucosal immunity that are necessary to interrupt the infection process.^1^

Though typically not required by regulators for product approval, VE against infection, regardless of symptoms, is commonly estimated. Strategies to identify infections in the absence of symptoms include weekly swab collections,^7^ cross-sectional testing at routine follow-up visits,^8^ and serological assays for evidence of non-vaccine induced immune responses.^9^ There is also interest in using these data to further estimate how well vaccines avert asymptomatic infections in isolation, given their importance in the control of transmission and relevance for understanding vaccine mechanisms.

The standard approach for evaluating vaccine efficacy against asymptomatic infection is to compare the risk of asymptomatic infection among vaccinated and unvaccinated individuals.^10–14^ In one COVID-19 vaccine trial, there was much attention paid to an estimate of this effect near zero, which some interpreted as poor vaccine performance.^11^ Yet these analyses can be difficult to interpret because they combine the effects of vaccine preventing asymptomatic infections with the effect of converting symptomatic to asymptomatic infections.^15,16^ In the extreme, a vaccine that effectively prevents symptoms but not infection could return a negative VE against asymptomatic infection despite the vaccine having not harmed any individual.

Here, we introduce a new estimand, VE against *naturally asymptomatic infection*, which describes the efficacy of a vaccine in preventing asymptomatic infections among individuals who would naturally (i.e., in the absence of vaccine) be asymptomatic. Unlike the estimand usually targeted, this approach excludes individuals for whom vaccination converts an otherwise symptomatic infection to an asymptomatic infection. This exclusion results in an estimand that more closely mirrors the interpretations of vaccine efficacy estimands against any infection and against symptomatic disease, making it more interpretable than the standard VE against asymptomatic infection. Using a causal principal stratification framework,^17,18^ we describe this estimand and assumptions under which it can be estimated. We apply these methods to a randomized trial of the COVID-19 mRNA-1273 vaccine^9^ and show how our approach can be generalized to analogous estimands for other endpoints by estimating vaccine efficacy against mild diarrhea in a rotavirus vaccine trial.^19^

## 2 Causal vaccine efficacy estimands

We consider a randomized trial design in which enrolled individuals are randomized to receive vaccine (*V* = 1 if vaccine; 0 if placebo) and are followed for any evidence of infection (*Y*_Inf_ = 1 if evidence of infection; 0 otherwise) and symptoms (*Y*_Sym_ = 1 if infection and symptoms; 0 otherwise). We also consider *X*, a vector of pre-randomization confounders needed to satisfy the assumptions listed below. Without loss of generality, we assume that *X* is discrete-valued; if not, sums in equations below can be replaced by appropriate integrals.

We define counterfactual outcomes for infection status under vaccine *v* (*Y*_Inf_(*v*)) and symptom status under vaccine *v* (*Y*_Sym_(*v*)). Under each exposure, individuals will either have symptomatic infection, asymptomatic infection, or no infection. Implicit in this notation is the single unit treatment value assumption (SUTVA), which states that there is only a single formulation of the vaccine and that there is no interference between individuals in the population. The former assumption is generally reasonable in well controlled studies of vaccine products, while the latter may be more dubious depending on the study design and epidemiology of transmission in the population of interest. We proceed assuming no interference but note that our methods may be generalized to allow interference.

We additionally assume that individuals cannot have symptoms without being infected (*Y*_Sym_(*v*) ≤ *Y*_Inf_(*v*)) and the vaccine cannot cause harm with respect to infection (*Y*_Inf_(1) ≤ *Y*_Inf_(0)) or developing symptoms (*Y*_Sym_(1) ≤ *Y*_Sym_(0)). Under these assumptions, we can define the six mutually exclusive and exhaustive basic principal strata^20^ based on an individual’s expected outcome with vaccine and with placebo (see Table 1 and Figure 1). We also highlight sets of principal strata that share the same outcome under placebo, which we refer to as the *naturally acquired* outcome (i.e., Naturally Symptomatic, Naturally Asymptomatic, or Naturally Never Infected). Finally, we define the Naturally Infected strata as the union of the Naturally Symptomatic and Naturally Asymptomatic sets.

**Table 1:** Principal strata based on counterfactual infection *Y*_Inf_(*v*) and symptom status *Y*_Sym_(*v*) under assumption of no vaccine harm. The Naturally Infected (Naturally Infected) strata is the union of the Naturally Symptomatic and the Naturally Asymptomatic strata.

| Combined strata | Basic principal stratum | $Y_{\text{Inf}}(0)$ | $Y_{\text{Sym}}(0)$ | $Y_{\text{Inf}}(1)$ | $Y_{\text{Sym}}(1)$ |
| --- | --- | --- | --- | --- | --- |
| Naturally Symptomatic | Always Symptomatic | 1 | 1 | 1 | 1 |
|  | Symptoms Prevented | 1 | 1 | 1 | 0 |
|  | Both Prevented | 1 | 1 | 0 | 0 |
| Naturally Asymptomatic | Always Asymptomatic | 1 | 0 | 1 | 0 |
|  | Asymptomatic Prevented | 1 | 0 | 0 | 0 |
|  | Never Infected | 0 | 0 | 0 | 0 |

**Figure 1:**
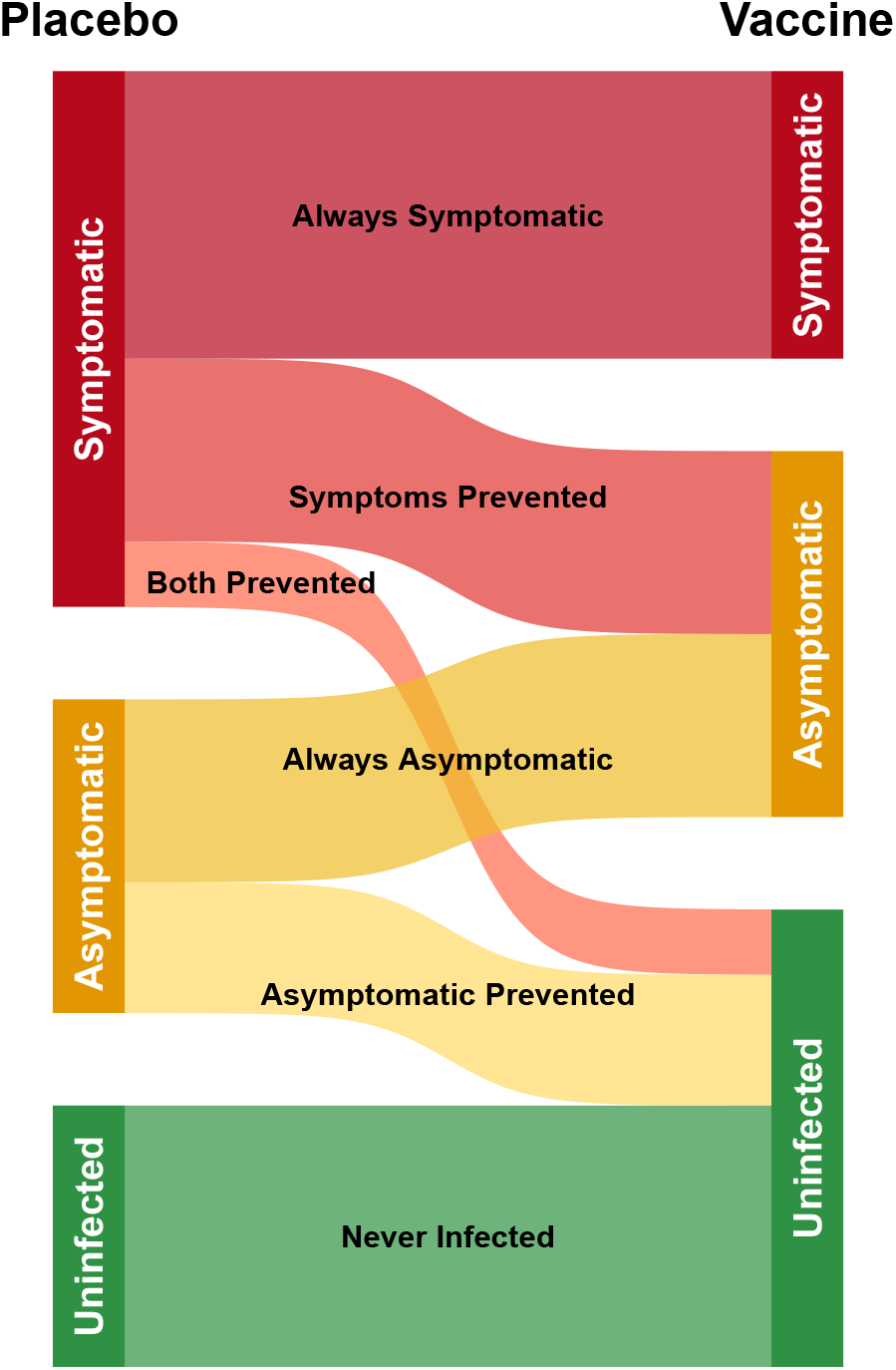
Visualization of six basic principal strata under the no harm assumption. The left side shows the counterfactual outcome under placebo (i.e., the *naturally acquired* outcome for individuals), the right side under vaccine. The potential outcomes are categorized as Symptomatic (*Y*_Inf_(*v*) = 1, *Y*_Sym_(*v*) = 1), Asymptomatic (*Y*_Inf_(*v*) = 1, *Y*_Sym_(*v*) = 0) and Uninfected (*Y*_Inf_ = 0, *Y*_Sym_ = 0).

In the next sections, we describe several VE estimands and interpret them in terms of these principal strata.

We start with the most common estimands that describe vaccine impact on symptomatic infection and on any infection, irrespective of symptoms. We then describe how the principal stratification framework can be used to identify shortcomings with the most common formulation of vaccine impact on asymptomatic infection and offer an improved estimand.

### 2.1 VE against symptomatic infection

VE against symptomatic infection is defined based on the relative risk of symptomatic infection among those receiving vaccine compared to placebo as

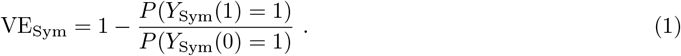

In terms of principal strata, this is

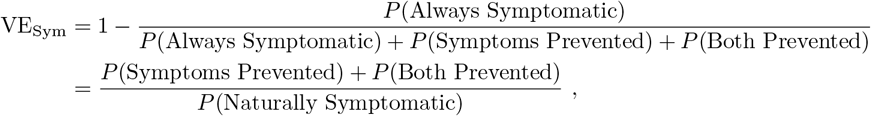

where e.g., *P* (Always Symptomatic) denotes the proportion of the population in the Always Symptomatic strata (i.e., the relative size of the Always Symptomatic strata in Figure 1). Writing VE_Sym_ in terms of principal strata demonstrates that it can be interpreted as the fraction of the Naturally Symptomatic strata that have their outcomes improved by the vaccine, either by having symptoms prevented or both symptoms and infection prevented (top left panel, Figure 2).

**Figure 2:**
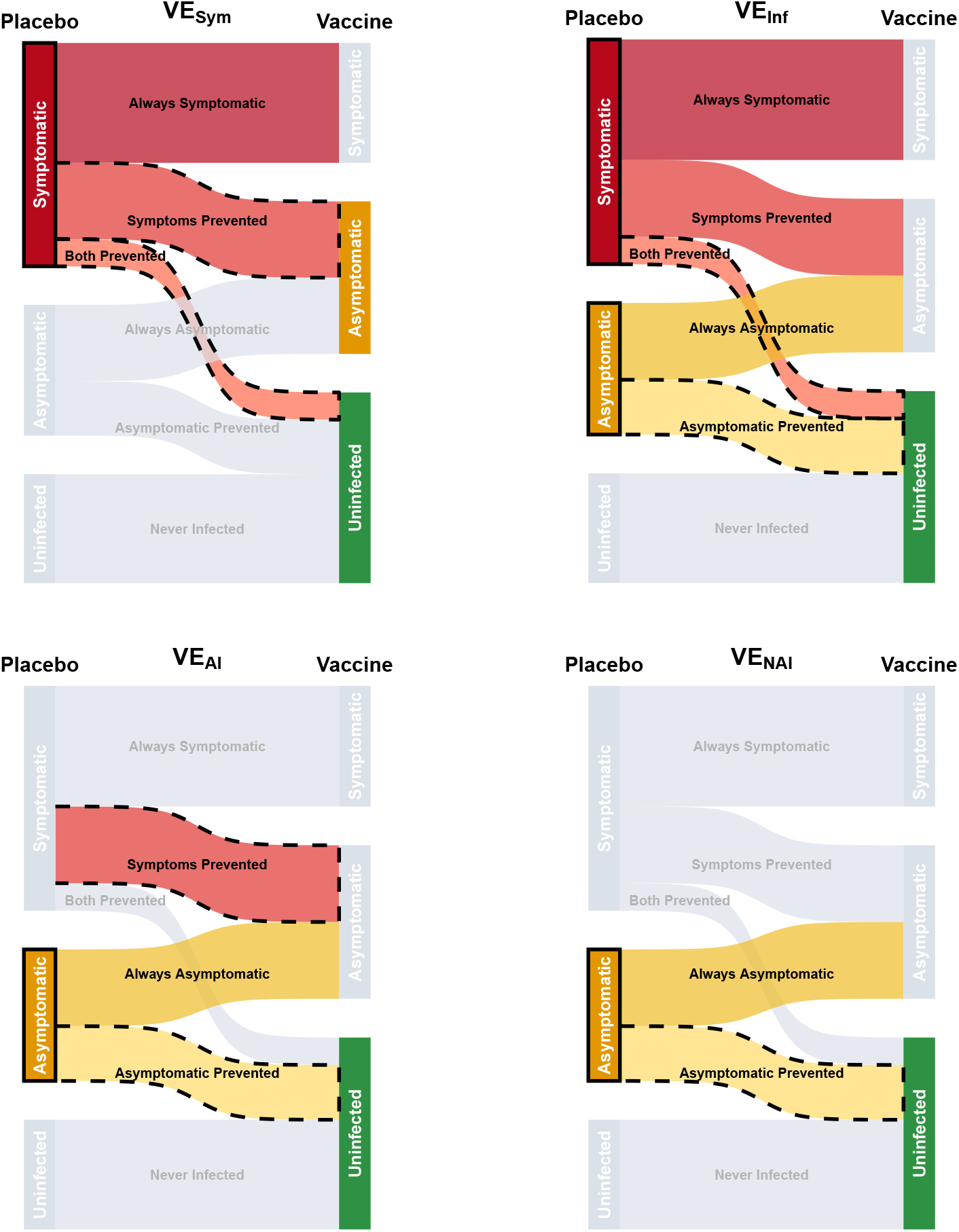
Vaccine efficacy parameters visualized in terms of principal strata. For each estimand, the denominator is comprised of the principal strata with the relevant naturally acquired outcome, highlighted by a solid black box. The principal strata that contribute to the numerator are highlighted via black dashed lines; strata that do not contribute to the estimand are shown in gray.

We could equivalently define the estimand as

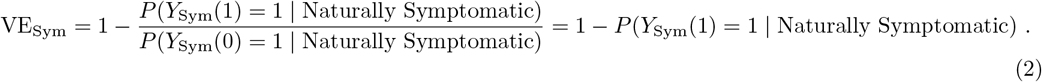

This formulation clarifies that those who are not Naturally Symptomatic (i.e., those with *Y*_Sym_(0) = 0) do not contribute to VE_Sym_ (gray boxes, top left panel, Figure 2); the estimand does not change when conditioning on *Y*_Sym_(0) = 1. It also provides an interpretation of VE_Sym_ as the risk reduction attributable to vaccine in the Naturally Symptomatic strata. This provides an analogous formulation to the novel VE_NAI_ proposed below.

### 2.2 VE against infection

VE against infection regardless of symptoms is defined based on the relative risk of infection among those receiving vaccine compared to placebo as

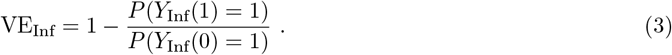

In terms of principal strata, this is

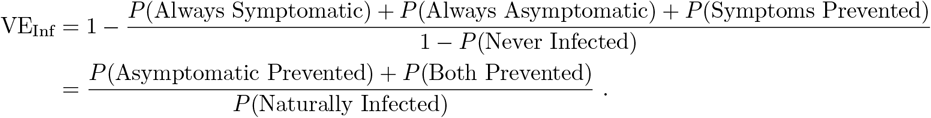

Thus, VE_Inf_ can be interpreted as the fraction of the Naturally Infected strata that either have their asymptomatic infection prevented or have both symptoms and asymptomatic infection prevented by the vaccine (top right panel, Figure 2). Equivalently,

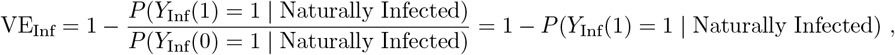

which again provides an interpretation as the risk reduction attributable to vaccine in the Naturally Infected subpopulation (i.e., those with *Y*_Inf_(0) = 1). Individuals that are immune to infection (*Y*_Inf_(0) = 0) do not contribute to the estimand.

### 2.3 VE against asymptomatic infection

A common, but often misinterpreted, way of quantifying VE against asymptomatic infections is by comparing the risk of asymptomatic infection between vaccinated and unvaccinated individuals as

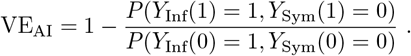

In terms of principal strata, this is

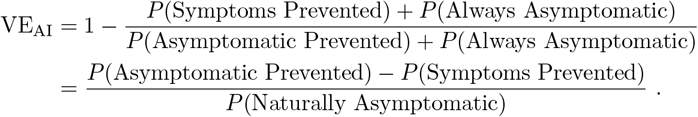

Since it combines vaccine effects of preventing infections and preventing symptoms, VE_AI_ is different than VE_Sym_ and VE_Inf_. While it can still be interpreted as a marginal change in the incidence of asymptomatic infections, it *cannot* be interpreted as the reduction in risk of an outcome among those who would naturally have that outcome (i.e., there is no equivalent conditional formulation as for the estimands above). While the denominator analogously defines the Naturally Asymptomatic population, the numerator is not a fraction of that population because individuals in the Symptoms Prevented stratum are Naturally *Symptomatic* (bottom left panel, Figure 2). Therefore, unlike VE_Sym_ and VE_Inf_ (and VE_NAI_ below), VE_AI_ is not bounded between 0 and 1 by the assumption of no vaccine harm.

### 2.4 VE against naturally asymptomatic infection

We propose an alternative estimand, VE against naturally asymptomatic infection, which is defined by comparing the risk of asymptomatic infections among individuals who would be asymptomatic under placebo:

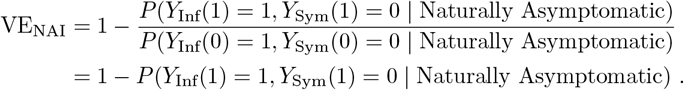

In terms of principal strata, this is

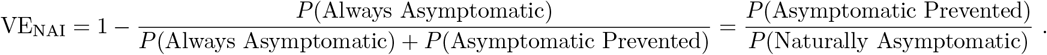

This estimand has a parallel interpretation to that of VE_Sym_ and VE_Inf_ as the fraction of the Naturally Asymptomatic strata that has their outcome improved by the vaccine (bottom right panel, Figure 2), and is bounded between 0 and 1 under the assumption of no vaccine harm.

However, there is an important difference between these estimands. VE_Sym_ and VE_Inf_ both have *unconditional* formulations (equations (1) and (3), respectively), implying that they are identifiable in the context of a randomized trial under minimal assumptions (e.g., comparing observed outcomes between randomized arms). On the other hand, there is no equivalent unconditional formulation for VE_NAI_, such that identification requires stronger assumptions in order to appropriately condition on the Naturally Asymptomatic strata.

We note that VE_NAI_ also generalizes to other hierarchies of endpoints. For example, vaccine efficacy against specifically mild disease was reported for COVID-19 vaccines.^12,21^ This estimand suffers from the same limitation as that for asymptomatic infection if the vaccine converts moderate/severe symptoms to mild. A VE parameter against naturally mild infections can be formulated by letting *Y*_Inf_ denote incidence of mild disease and *Y*_Sym_ denote incidence of moderate-to-severe disease (see eAppendix D for example in a rotavirus vaccine trial).

## 3 Identification and estimation of VE_NAI_

### 3.1 Point identification

VE_NAI_ can be point identified from observed data under the assumption of no vaccine harm and the following sufficient set of assumptions:

(A1) *No unmeasured confounding of infection risk and vaccine receipt* : *Y*_Inf_(0) ⊥ *V* | *X* and *Y*_Inf_(1) ⊥ *V* | *X*
(A2) *No unmeasured confounding of symptoms under placebo and vaccine receipt* : *Y*_Sym_(0) ⊥*V* | *Y*_Inf_(0) = 1, *X*
(A3) *Positivity of vaccine assignment* : 0 *< P* (*V* = 1 | *X* = *x*) *<* 1 for all *x*
(A4) *No unmeasured confounding of symptoms under placebo and infection under vaccine among the Naturally Infected* : *Y*_Sym_(0) ⊥ *Y*_Inf_(1) | *Y*_Inf_(0) = 1, *X*

The first three assumptions are satisfied by design in a randomized trial. Otherwise, these assumptions require *X* to be sufficient to control confounding of vaccine status and both infection and symptoms, and to satisfy positivity condition (A3). Assumption (A4) is a form of principal ignorability assumption^22,23^ that requires control for any common causes of having symptoms under placebo and being infected under vaccine. Risk factors for exposure, markers of host susceptibility (e.g., age), and pre-existing pathogen-specific or cross-protective immunity are candidates for *X* that may be capable of satisfying this assumption. Due to the fundamentally cross-world and therefore untestable nature of this assumption, we describe the estimation of bounds without this assumption and a sensitivity analysis in the following sections.

Under the assumptions described above, VE_NAI_ can be point-identified by

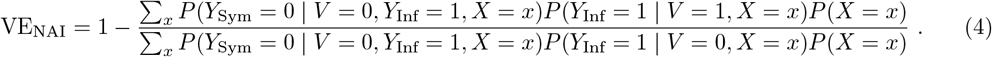

A proof is included in eAppendix A. This identification result is similar to that for VE_AI_,

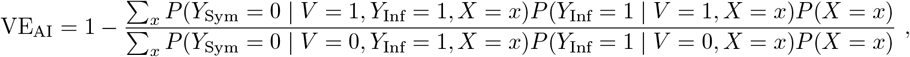

where the only difference is the first term in the numerator of the risk ratio. In VE_AI_, the sum in the numerator describes a stratum-specific probability of being infected among the vaccinated (*P* (*Y*_Inf_ = 1 |*V* = 1, *X* = *x*)) multiplied by the probability of not developing symptoms given infection among the vaccinated (*P* (*Y*_Sym_ = 0 | <u>*V* = 1</u>, *Y*_Inf_ = 1, *X* = *x*)). However, the observed asymptomatically infected vaccinated individuals are a combination of Always Asymptomatic and Symptoms Prevented, whereas we wish to isolate only the Always Asymptomatic for identification of VE_NAI_. To do this, we “impute” the fraction of vaccinated infecteds who are Naturally Asymptomatic (i.e., the Always Asymptomatic) based on the fraction of the infected placebo participants that are asymptomatic, *P* (*Y*_Sym_ = 0 | <u>*V* = 0</u>, *Y*_Inf_ = 1, *X* = *x*).

This identification result gives additional intuition into (A4) – the assumption requires that conditional on *X*, infected vaccinated participants are not systematically different from infected placebo recipients with respect to their likelihood of naturally developing symptoms (eFigure 4). Another implication of (A4) is that within strata of *X*, VE_NAI_ = VE_Inf_. Thus, under this assumption VE_NAI_ can be formulated as a weighted average of stratum-specific VE_Inf_ (see eAppendix B for proof and discussion).

### 3.2 Bounds on VE_NAI_

Bounds on VE_NAI_ can be derived by recognizing that outcomes directly observed in a trial partially identify (combinations of) principal strata. For example, from Figure 1, we can infer that vaccinated participants who are observed to be symptomatic must be in the Always Symptomatic stratum and thus *P* (Always Symptomatic) = *P* (*Y*_*I*_ = 1, *Y*_*S*_ = 1 |*V* = 1). Similarly, asymptomatic placebo recipients must be in either the Always Asymptomatic or Asymptomatic Prevented strata and thus *P* (Always Asymptomatic)+ *P* (Asymptomatic Prevented) = *P* (*Y*_*I*_ = 1, *Y*_*S*_ = 0, | *V* = 0). This logic can be used to derive bounds without making assumption (A4).

First, VE_AI_ is a lower bound for VE_NAI_, since

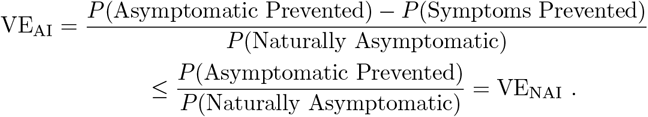

Because VE_AI_ has an unconditional formulation, it can be identified by comparing the proportion of asymptomatic infections between vaccinated and unvaccinated individuals, resulting in the following lower bound:

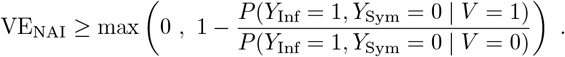

While VE_AI_ can be negative, we do not allow this bound to be *<* 0 to be consistent with the no harm assumption. This lower bound can be described as assuming the “worst-case” scenario for VE_NAI_ – that the vaccine converts no individuals from symptomatic to asymptomatic infection (*P* (Symptoms Prevented) = 0).

An upper bound can be derived by noting that the proportion of the population that is either Asymptomatic Prevented or Both Prevented can be identified by

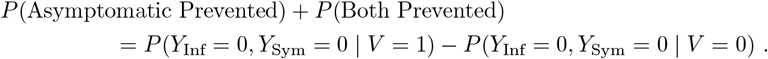

This is true since the observed uninfected vaccine recipients must be either Never Infected, Both Prevented, or Asymptomatic Prevented, while observed uninfected placebo recipients must be Never Infected (Figure 1). The difference gives the combined size of the Both Prevented and Asymptomatic Prevented strata. Therefore, we can derive an upper bound by assuming a “best-case” scenario for the vaccine – that there are no individuals in the P(Both Prevented) stratum. In this scenario, VE_NAI_ can be directly identified based on the observed data because the difference in uninfected individuals between arms must equal the Asymptomatic Prevented (numerator of VE_NAI_), and the observed infected placebo recipients are the Naturally Asymptomatic (denominator of VE_NAI_). Specifically, *P* (Asymptomatic Prevented) = *P* (*Y*_Inf_ = 0, *Y*_Sym_ = 0 | *V* = 1) − *P* (*Y*_Inf_ = 0, *Y*_Sym_ = 0 | *V* = 0), while *P* (Naturally Asymptomatic) = *P* (*Y*_Inf_ = 1, *Y*_Sym_ = 0 | *V* = 0).

An upper bound on VE_NAI_ is therefore

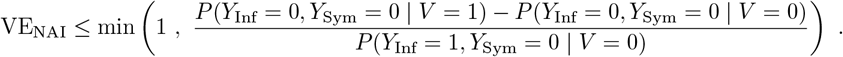

Intuitively, this upper bound would occur if the total vaccine effect on infection occurred only in the Naturally Asymptomatic strata.

### 3.3 Sensitivity analysis

Given the untestable nature of (A4), we propose a sensitivity analysis to evaluate robustness to this assumption. Similar sensitivity analyses have been proposed for other principal strata estimands.^23^ (A4) implies that conditional on *X*, the probability of being asymptomatic under placebo is the same among the Naturally Infected for whom vaccine does not prevent infection (the Always Symptomatic, Always Asymptomatic, Symptoms Prevented strata) and the Naturally Infected for whom vaccine prevents infection (the Asymptomatic Prevented and Both Prevented strata):

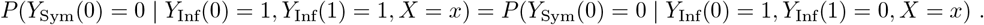

We may generalize this assumption by instead assuming that these probabilities are proportional by *δ*:

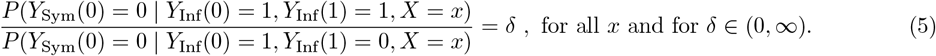

In situations in which it may be reasonable to assume different values for *δ* based on different covariate profiles, the ratio in (5) could be allowed to vary with *x*.

Under (5), we can identify VE_NAI_ by

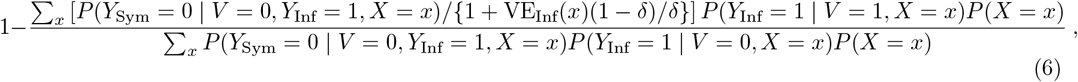

where VE_Inf_(*x*) is the observed data stratum-specific VE against any infection regardless of symptoms, VE_Inf_(*x*) = 1 − *P* (*Y*_Inf_ = 1 | *V* = 1, *X* = *x*)*/P* (*Y*_Inf_ = 1 | *V* = 0, *X* = *x*) (see proof in eAppendix A).

A sensitivity analysis can study how VE_NAI_ varies as a function of *δ*. Evidence of efficacy against naturally asymptomatic infections would be strengthened if VE_NAI_ *>* 0% for all scientifically plausible values of *δ*.

In practice, it is likely reasonable to assume that *δ <* 1 for many vaccines. For example, for SARS-CoV-2, humoral immunity is primarily responsible for preventing infection, while cellular immunity may be necessary to reduce symptom severity.^24^ Because both of these responses are components of the adaptive immune response, they are likely to be positively correlated – individuals who have strong humoral immune responses tend to have strong cellular immune responses as well. Thus, we expect that individuals who become infected even after they receive vaccine (i.e., those in the numerator of (5)) are those who have poor antibody responses and therefore likely have poor cellular responses too, leading to relatively high rates of symptom onset following infection. On the other hand, individuals who are protected by the vaccine (i.e., those in the denominator) are expected to have relatively stronger antibody responses and therefore likely have cellular immune responses that are at least as strong as those in the numerator. If this is true, such that *δ <* 1, the point identification formula (4) will lead to *conservative* estimates of VE_NAI_, as the value of the sensitivity parameter (6) increases as *δ* decreases.

Another potentially valuable approach to sensitivity analysis would be to determine the value of *δ* such that VE_NAI_ = VE_AI_ (i.e., the lower bound) and evaluate whether that value is scientifically plausible. In line with the rationale above, the larger this value of *δ*, the more implausible it is, suggesting rather that VE_NAI_ *>* VE_AI_. Such an analysis could provide quantitative evidence that VE_NAI_ is larger than VE_AI_, without claiming to provide an exact estimate of its value.

### 3.4 Point estimation of VE_NAI_

Estimation of VE_NAI_ and related sensitivity parameters can be based on (i) 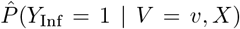, an estimate of the conditional cumulative risk of any infection over the study period under vaccine (*v* = 1) and placebo (*v* = 0) and (ii) 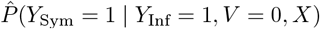, an estimate of the conditional probability of being symptomatic given infection under placebo. The former could be based on logistic regression models if there is no right censoring or appropriate survival analysis methods if there is right censoring (see eAppendix C for details on estimation using Cox proportional hazard regression models). The estimate of the probability of being symptomatic given infection could be based on logistic regression in infected placebo recipients. An estimator of VE_NAI_ is then

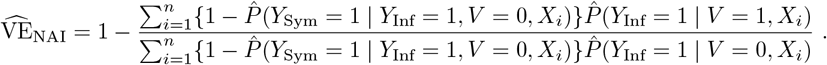

Similarly, the sensitivity analysis can be conducted by plugging in estimated quantities to equation (6). Standard errors, confidence intervals and hypothesis tests for both the point identification, as well as the sensitivity analysis can be based on the nonparametric bootstrap.

## 4 Application

### 4.1 The COVE study

The COVE study (NCT04470427) was a Phase 3 randomized placebo-controlled trial of the mRNA-1273 SARS-CoV-2 vaccine that enrolled individuals in the United States from July-October 2020.^9^ A total of 30,420 adults at risk of SARS-CoV-2 infection or severe COVID-19 disease with no known prior SARS-CoV-2 infections were enrolled and randomized 1:1 to receive two doses of the mRNA-1273 vaccine or placebo 28 days apart. Participants were followed for symptomatic COVID-19, defined by positive PCR test and presence of at least two symptoms. Asymptomatic SARS-CoV-2 infections were identified by either a positive PCR test without symptoms or seroconversion at day 57 or at the end of blinded follow-up.^10^ Our analysis matches the original primary analysis which included outcomes with onset at least 14 days after the second dose in the subset of the per-protocol population (i.e., those who received two doses) who were seronegative at baseline. We analyzed infection and disease endpoints observed through the end of blinded follow-up, for which previous estimates of VE_AI_ were considerably lower than VE_Sym_ and VE_Inf_.^10^

### 4.2 Analysis methods

We estimated the cumulative incidence of infection regardless of severity using two cause-specific Cox proportional hazards regression models (see eAppendix C for details). One model was used to estimate the hazard for symptomatic infections, which were ascertained continuously in time via passive monitoring for symptomatic COVID-19 infections; the second model was used to estimate the hazard for asymptomatic infections, which were ascertained at two fixed time points following vaccination. The probability for developing symptoms given infection was estimated using logistic regression. All regression models were adjusted for age, sex, an indicator for being at high risk for severe COVID-19, and race/ethnicity. We generated 95% confidence intervals for the various vaccine efficacy parameters based on percentiles of 1000 nonparametric bootstrap estimates.

### 4.3 Results

The per-protocol population included 14,254 individuals who received vaccine and 14,069 individuals who received placebo. There were 66 symptomatic cases of COVID-19 among those receiving vaccine and 841 among those receiving placebo. Asymptomatic infections were identified in 214 and 498 individuals receiving vaccine and placebo, respectively. At 200 days of follow-up, the efficacy of mRNA-1273 against symptomatic infection was 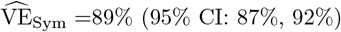 and against any infection was 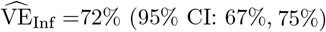. The naive estimate of efficacy against asymptomatic infection was 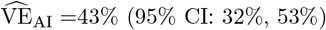. The upper bound on VE_NAI_ was uninformative as it was estimated to be 1, which is not surprising given the vaccine’s strong protective efficacy against symptomatic infection. Thus, the analysis suggests that without the cross-world independence assumption (A4), we estimate that 43% *<* VE_NAI_ *<* 100%. If we assume (A4), we obtain an estimate of 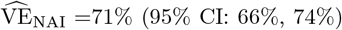, indicating much higher efficacy against naturally asymptomatic infections than is suggested by VE_AI_.

In our sensitivity analysis, we determined the value of *δ* such that VE_NAI_ equaled the assumption-free lower bound of 43% (*δ* ≈ 3.0). In order for VE_NAI_ to equal this value, individuals who would be infected despite vaccination must be around three times as likely to remain symptom free following infection when compared to individuals in whom the vaccine prevented infection. This is highly unlikely given our understanding of COVID-19 vaccines and immunology. Thus, our analysis provides evidence that the mRNA1273 vaccine likely converted symptomatic infections to asymptomatic infections for some individuals in the COVE trial. Allowing *δ* to range from 1*/*3 to 3, our sensitivity analysis showed estimates of VE_NAI_ ranging from 89%-43% (Figure 3) indicating that the vaccine’s ability to prevent naturally asymptomatic infections could be considerably higher than that implied not only by VE_AI_ but also the point identification of VE_NAI_.

**Figure 3:**
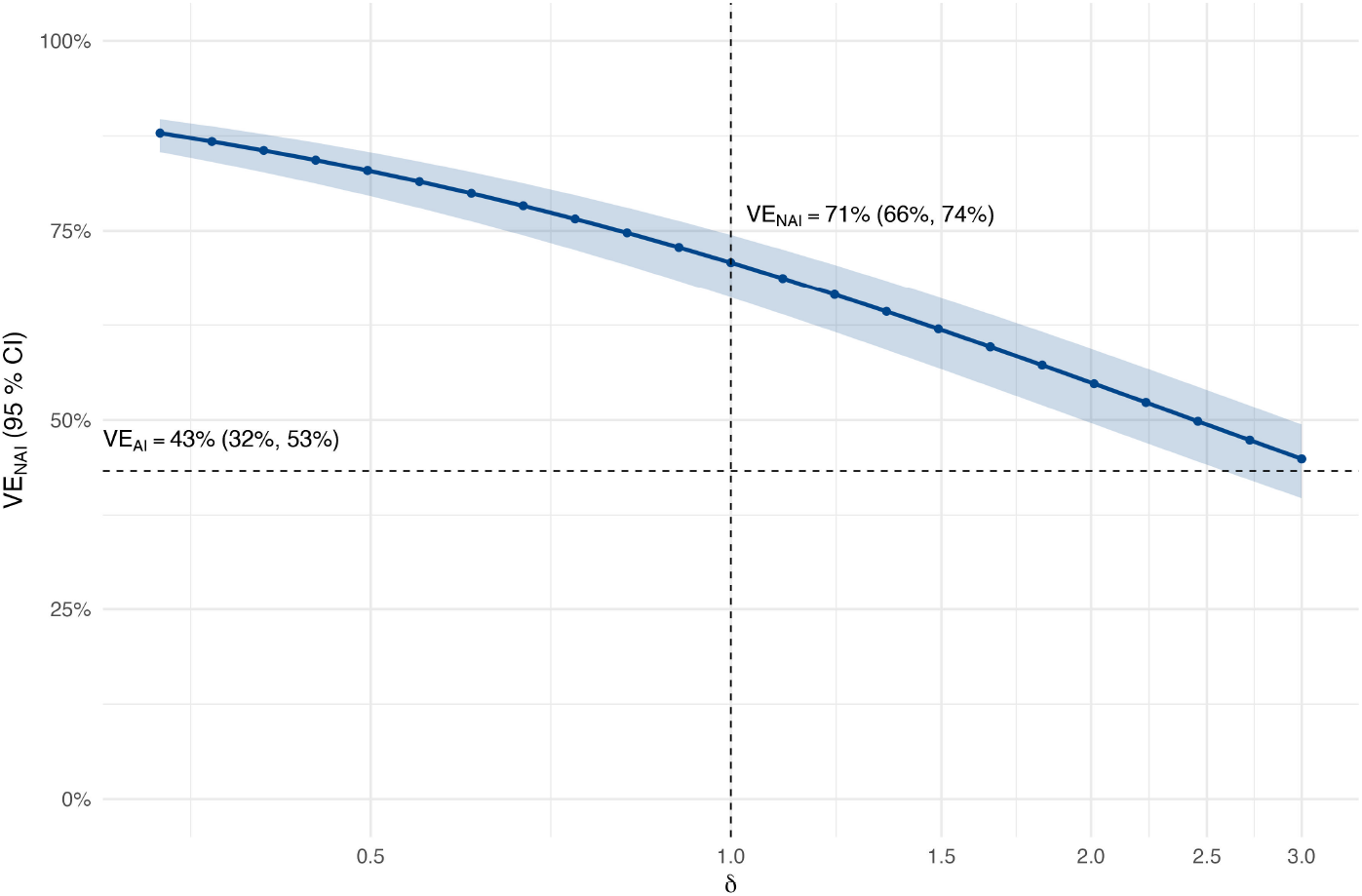
Sensitivity analysis of VE_NAI_. The horizontal dashed line indicates the estimate of VE_AI_, and the vertical dashed line indicates the estimate of VE_NAI_ under Assumption (A4).

## 5 Discussion

Our novel estimand, VE_NAI_, is analogous to other commonly reported vaccine effects (VE_Sym_ and VE_Inf_) and provides a more interpretable estimate of the impact of vaccine on asymptomatic infections than the commonly reported VE_AI_. Intuitively, VE_NAI_ can be interpreted as the reduction in risk of asymptomatic infections among those who would be asymptomatic in the absence of vaccine. Similar interpretations apply to other common VE estimands quantifying for example vaccine impacts on severe outcomes or death.

Our re-analysis of COVE demonstrated that VE_NAI_ for the mRNA-1273 vaccine was likely considerably larger in magnitude than the previously reported VE_AI_.^10^ Understanding and accurately characterizing impacts of vaccination on asymptomatic infections may be particularly important for pediatric studies of COVID-19 vaccines, where VE_AI_ was reported to be close to zero for both the ChAdOx1 nCoV-19 vaccine (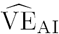 = 22%, 95% CI: -10%, 45%)^11^ and the mRNA-1273 vaccine (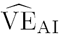 = 23%, 95% CI: -20%, 49% in children aged 2-5 years and 4%, 95% CI: -112%, 53% in children 6-23 months).^14^ Presentation of these results without a careful discussion of how the estimates are affected by individuals who have their symptoms but not the underlying infection prevented may lead to confusion and present challenges to conveying the benefits of vaccination, particularly since children are more likely to be Naturally Asymptomatic than adults.

The estimands explored here are all relative effects, mirroring the most commonly reported effects in vaccine trials. However, additive effects (i.e., risk differences) may in some cases be more relevant to public health decision-making. Importantly, unlike their relative effect counterparts, the additive versions of VE_Sym_ and VE_Inf_ do not have an equivalent formulation that is conditional on the Naturally Symptomatic or Naturally Infected respectively. The additive version of VE_AI_ is therefore not discrepant with the other effects, and when effects on the total burden of asymptomatic infections are of primary interest, an estimate of VE_AI_ on the additive scale may be most informative. Specifically, decisions about interventions aimed at preventing transmission from asymptomatic infections should be informed by the total number of expected asymptomatic infections, not only those among the Naturally Asymptomatic. Evidence that vaccine increases the total number of asymptomatic infections (i.e., VE_AI_ *<* 0) could be used to help justify recommendations for additional preventive measures (e.g., masking or social distancing) in vaccinated individuals.

The proposed methods are limited by the strong cross-world assumption required for point identification and the potential for uninformative bounds. For regulatory purposes (e.g., if asymptomatic infections are included as a secondary or exploratory endpoint), an informative lower bound is likely more important than an informative upper bound, potentially ruling out a lack of efficacy against asymptomatic infections.

Unfortunately, the settings in which *P* (Symptoms Prevented) may be relatively large (which would result in a non-informative lower bound) are precisely the settings in which we might expect VE_AI_ *<* 0 and therefore VE_NAI_ may be of considerable interest. Thus, it may be insufficient in many practical settings to only consider bounds for VE_NAI_ without making further assumptions. Our proposed sensitivity analyses provide means of exploring the robustness of VE_NAI_ to the cross-world assumption and potentially provide more realistic, informative upper bounds based on plausible values of *δ*.

The impact of the cross-world independence assumption may also be mitigated by exploring separable causal effects formulations of these estimands. Such approaches have been used in mediation problems that rely on cross-world assumptions^25^ and may be appealing given the potential for distinct immune mechanisms to be responsible for blocking infections versus symptoms.

Our approach can also be used to estimate other types of vaccine effects, including a causal analogue to VE for progression (see eAppendix E for details). It is also of interest to develop more robust estimators of VE_NAI_ and related quantities, e.g., based on cross-fitted machine learning-based regression models. We expect that given the randomized trial context, these estimators would enjoy desirable robustness properties to some patterns of misspecification of the regression models.

Reporting VE_NAI_ in future trials will help equip practitioners to answer questions from the public such as, “what is the benefit of getting vaccinated if I would have been asymptomatic, anyway?” Alongside other vaccine effects, VE_NAI_ would improve interpretability of results, could broaden understanding of vaccine impact on transmission, and provide insights into immunological mechanisms.

## Data Availability

Data from the COVE study are available on reasonable request to the corresponding author. Data from the PROVIDE study are publicly available at clinepidb.org. Computing code for the analysis is available at http://github.com/allicodi/ve_nai and for a corresponding R package at https://github.com/allicodi/VEnai.

https://clinepidb.org/

https://github.com/allicodi/VEnai

## Acknowledgments

We thank the volunteers who participated in the PROVIDE and COVE trials and the PROVIDE study team including Beth Kirkpatrick, Rashidul Haque, and William A Petri, Jr.

## eAppendix A: Proofs

The full list of required assumptions can be restated as follows:

(A0.0) *Causal consistency. Y*_Inf_ = *V × Y*_Inf_(1) +(1 − *V* ) *× Y*_Inf_(0) and *Y*_Sym_ = *V × Y*_Sym_(1) +(1 − *V* ) *× Y*_Sym_(0). (A0.1) *No symptomatic infection without infection*: *Y*_Sym_(*v*) ≤ *Y*_Inf_(*v*);
(A0.2) *No vaccine harm with respect to infection nor developing symptoms*: *Y*_Inf_(1) ≤ *Y*_Inf_(0) and *Y*_Sym_(1) ≤ *Y*_Sym_(0);
(A1) *No unmeasured confounding of infection risk and vaccine receipt* : *Y*_Inf_(0) ⊥ *V* | *X* and *Y*_Inf_(1) ⊥ *V* | *X*
(A2) *No unmeasured confounding of symptomatic infection risk and vaccine receipt* : *Y*_Sym_(0) ⊥*V* | *Y*_Inf_(0) = 1, *X*
(A3) *Positivity of vaccine assignment* : 0 *< P* (*V* = 1 | *X* = *x*) *<* 1 for all *x*.
(A4) *No unmeasured confounding of symptomatic infection under placebo and infection under vaccine*: *Y*_Sym_(0) ⊥ *Y*_Inf_(1) | *Y*_Inf_(0) = 1, *X*

### A1. Proof for identification result for VE_NAI_

*Proof*. By definition, VE_NAI_ = 1 − *P* (*Y*_Inf_(1) = 1, *Y*_Sym_(1) = 0 | Naturally Asymptomatic). Due to (A0.2), *P* (*Y*_Inf_(1) = 1, *Y*_Sym_(1) = 0| Naturally Asymptomatic) = *P* (*Y*_Inf_(1) = 1| *Y*_Inf_(0) = 1, *Y*_Sym_(0) = 0). Then we can write

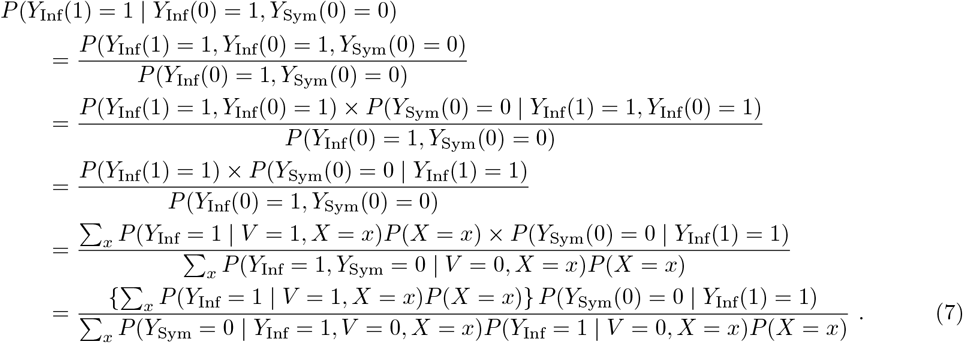

The first and second equalities follow from definitions of conditional probability. The third equality follows from (A0.2). The fourth equality follows from (A1)-(A2). The final equality follows from laws of conditional probability. Next, we can write

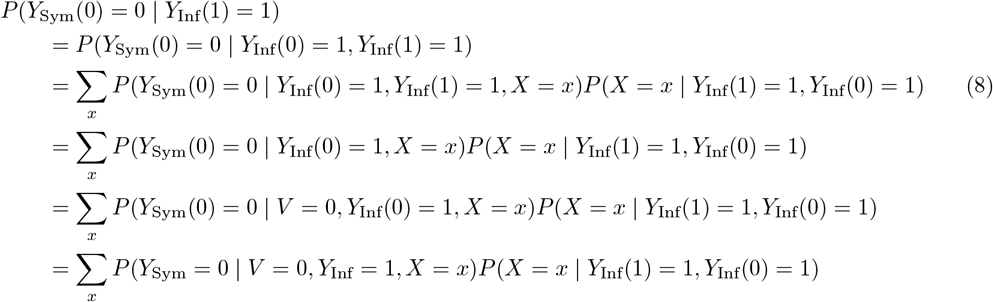

The first equality follows from (A0.1) and (A0.2), the second from the law of total probability. The third follows from (A4), the fourth from (A2). The final equality follows from (A0.0). Next, we consider *P* (*X* | *Y*_Inf_(1) = 1, *Y*_Inf_(0) = 1):

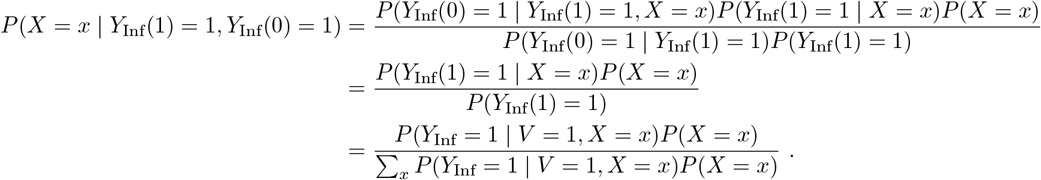

The first equality follows from Bayes Theorem, the second from (A0.1) and (A0.2). The third follows from (A1). Finally, assumptions (A0.2) and (A3) ensure that all conditional probabilities are well defined for each term in the sum. Thus, we have shown that the numerator of (7) is

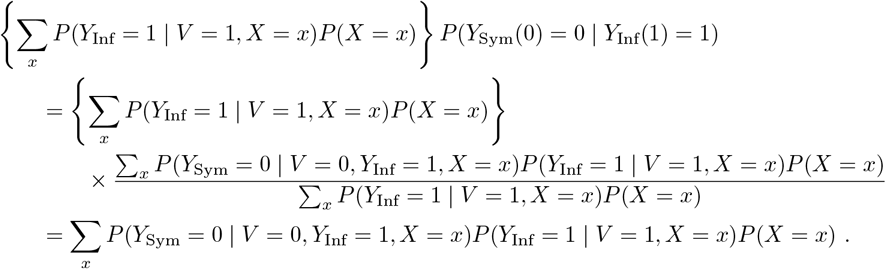

Thus, we have that

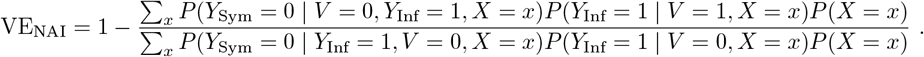

□

### A2. Proof for identification result for sensitivity analysis on VE_NAI_

*Proof*. Identification proceeds exactly as that above through equation (8). Identification of *P* (*X* = *x* | *Y*_Inf_ = 1, *Y*_Inf_(0) = 1) is also identical to the proof above. Thus, it only remains to provide an alternative identification for *P* (*Y*_Sym_(0) = 0 | *Y*_Inf_(0) = 1, *Y*_Inf_(1) = 1, *X* = *x*). Using the law of total probability and (5), we have

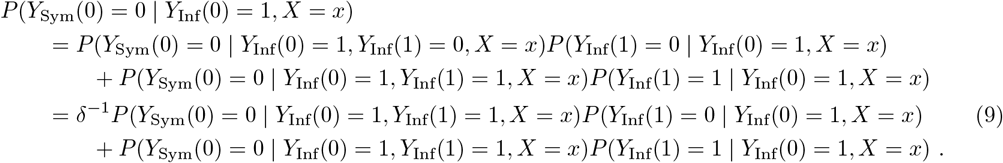

Bayes’ Theorem and Assumptions (A0.1) and (A0.2) imply that

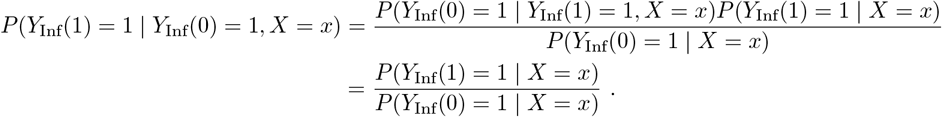

Rearranging terms in (9), we have that

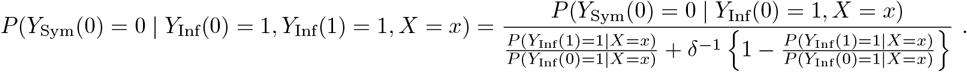

The numerator is identified under (A2) by *P* (*Y*_Sym_ = 0| *V* = 0, *Y*_Inf_ = 1, *X* = *x*). The denominator is identified under (A1) by

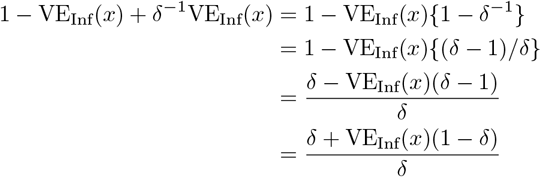

□

### A3. Visual representation of (A4)

Assumption (A4) is visually represented in Figure 4, in which proportions are depicted by outlining components of the numerator with dashed black lines and components of the denominator with solid black boxes. The assumption states that within each covariate strata, the fraction of the Naturally Infected who are Naturally Asymptomatic (depicted in the left panel) is the same as the fraction of infected vaccinated individuals that are Naturally Asymptomatic (i.e., in the Always Asymptomatic stratum; depicted in the right panel). The corresponding distributions of principal strata are shown in Table 2. The left-most column can be used to confirm that

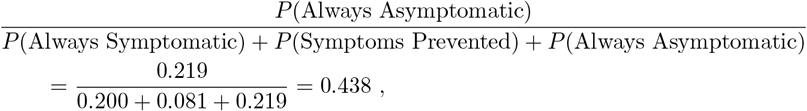

is the same as

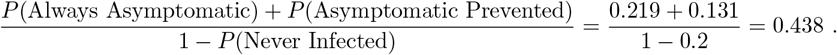

**Table 2:** Three hypothetical settings where (A4) is satisfied (left column), where (A4) is violated such that *δ <* 1 (right column), and where (A4) is violated such that *δ >* 1. VE_NAI_ (obs.) represents the value that would be obtained using the identifying formula based on (A4).

|  | Figure 4 | Figure 5 | Figure 6 |
| --- | --- | --- | --- |
| $P(\text{Always Symptomatic})$ | 0.200 | 0.100 | 0.200 |
| $P(\text{Symptoms Prevented})$ | 0.081 | 0.281 | 0.031 |
| $P(\text{Both Prevented})$ | 0.169 | 0.019 | 0.269 |
| $P(\text{Always Asymptomatic})$ | 0.219 | 0.069 | 0.319 |
| $P(\text{Asymptomatic Prevented})$ | 0.131 | 0.231 | 0.031 |
| $P(\text{Never Infected})$ | 0.200 | 0.300 | 0.150 |
| $\text{VE}_{\text{Inf}}$ | 37.5% | 35.7% | 35.3% |
| $\text{VE}_{\text{Sym}}$ | 55.6% | 75.0% | 60.0% |
| $\text{VE}_{\text{AI}}$ | 14.3% | -16.7% | 0.0% |
| $\delta$ | 1.000 | 0.165 | 5.564 |
| $\text{VE}_{\text{NAI}} (\text{true})$ | 37.5% | 77.1% | 8.9% |
| $\text{VE}_{\text{NAI}} (\text{obs.})$ | 37.5% | 35.7% | 35.3% |

While the figure displays a single hypothetical covariate strata, the assumption should hold in all strata defined by covariates *X*, and need not hold marginally. The value of the ratio is allowed to vary by covariate strata (i.e., certain strata may have a greater or lesser proportion of Naturally Infected who are Naturally Asymptomatic); however, the ratio should be the same on the left side as the right side of the figure.

We can also consider how this figure would look when (A4) is not satisfied. For example, we can consider the setting in Figure 6, with corresponding principal strata distributions shown in the middle column of Table 2. In this case, we can confirm that the true fraction of Always Asymptomatic among those who would be infected under vaccine is

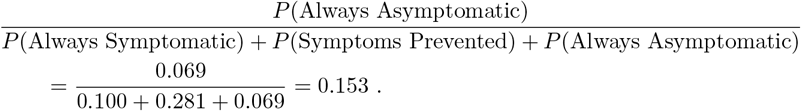

**Figure 4:**
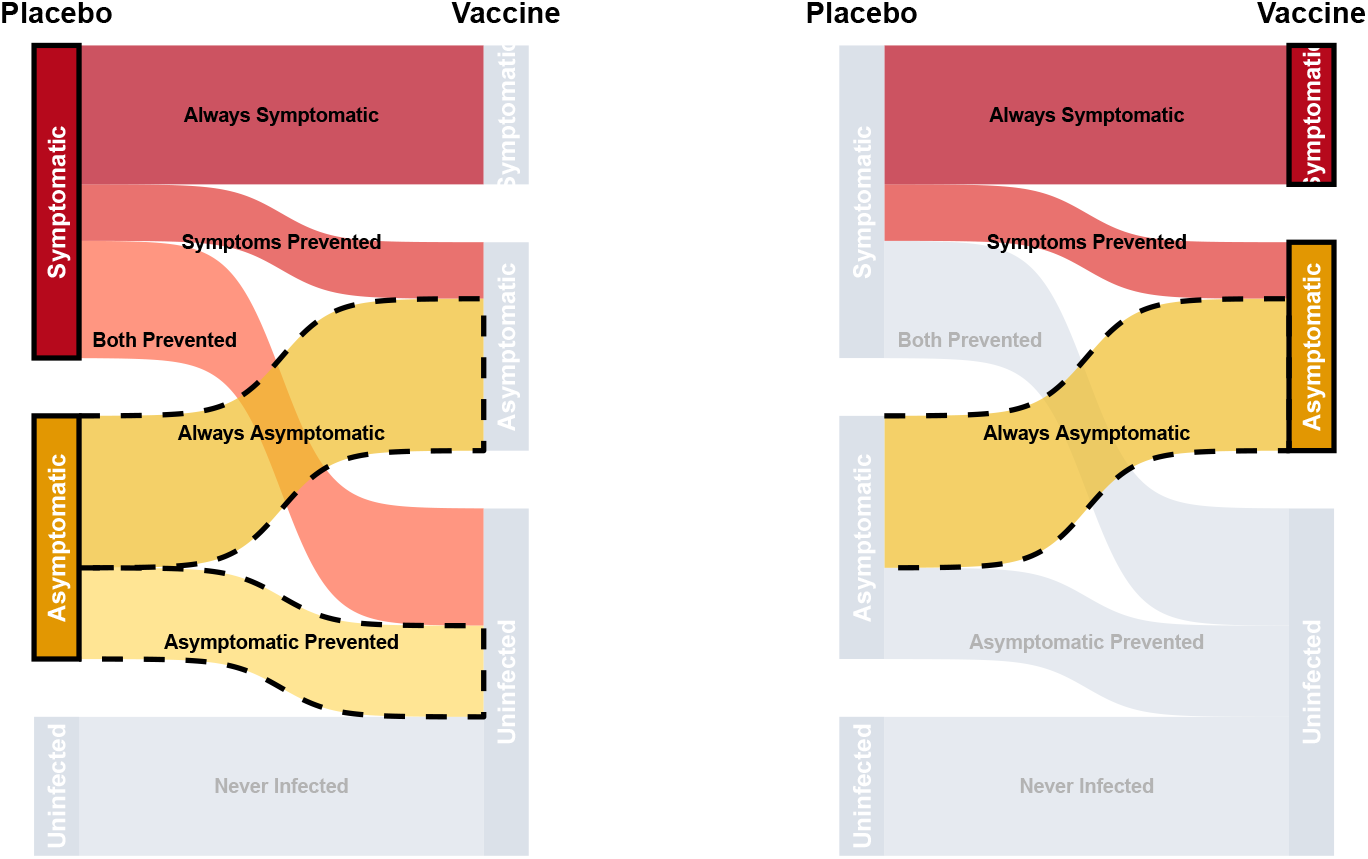
Visual depiction of assumption (A4). The fraction of the Naturally Infected (left panel, black boxed nodes) that are Naturally Asymptomatic (left panel, dashed flows) is equal to the fraction of the vaccinated infected (right panel, black boxed nodes) that are Always Asymptomatic (right panel, dashed flows).

**Figure 5:**
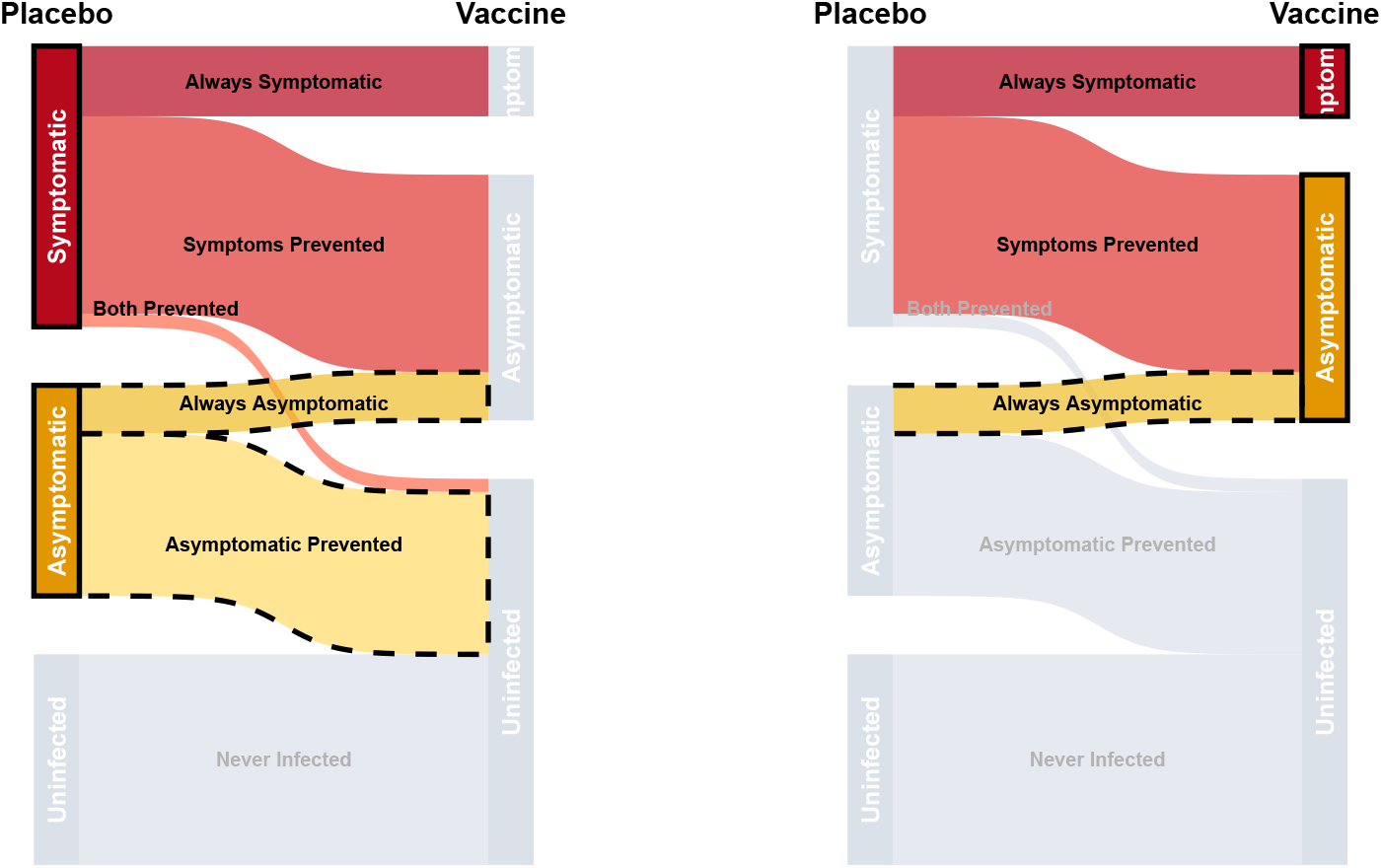
Visual depiction of a violation of assumption (A4) where *δ* = 0.165. The fraction of the Naturally Infected (left panel, black boxed nodes) that are Naturally Symptomatic (left panel, dashed flows) is larger than the fraction of the vaccinated infected (right panel, black boxed nodes) that are Always Asymptomatic (right panel, dashed flows).

**Figure 6:**
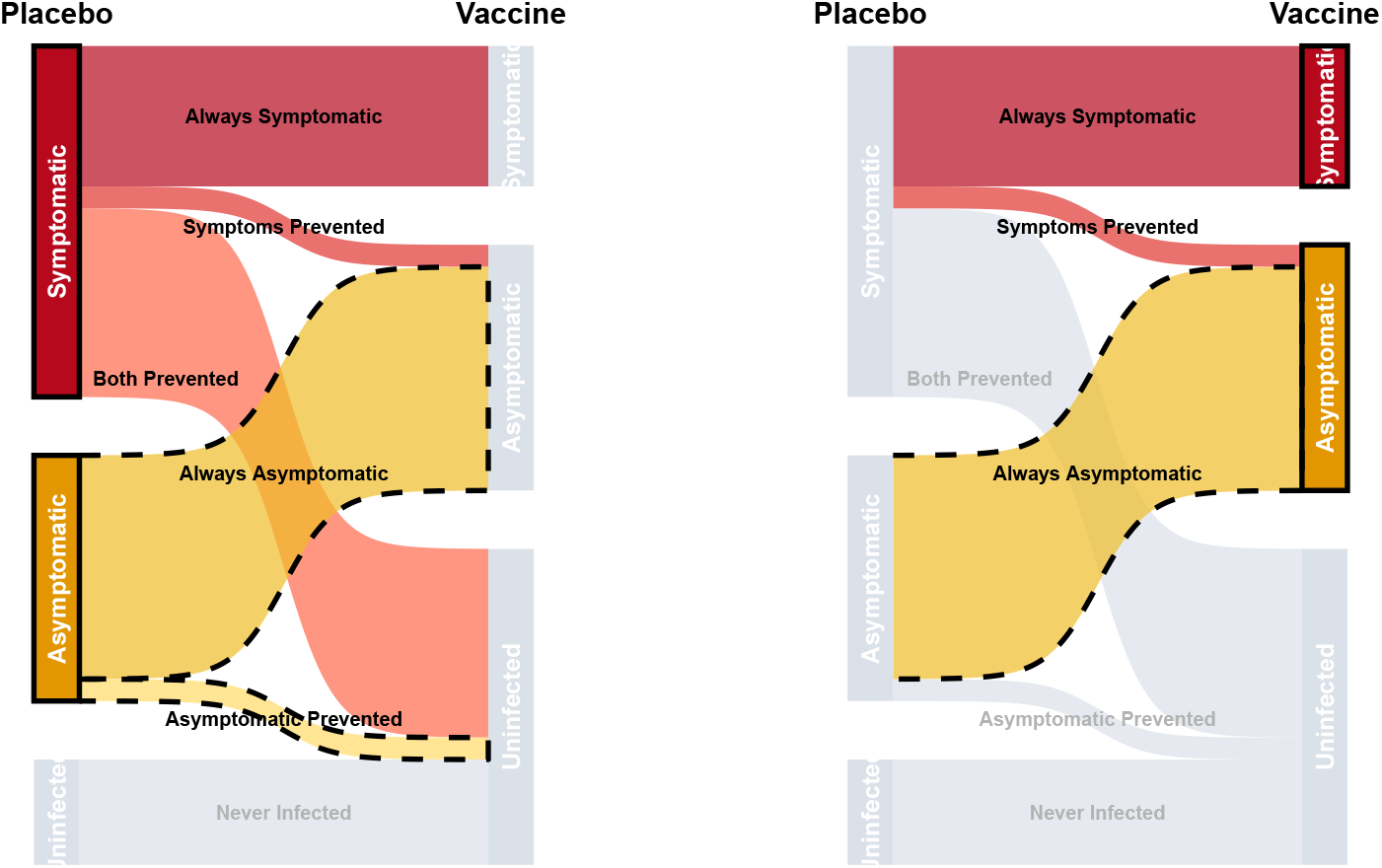
Visual depiction of a violation of assumption (A4) where *δ* = 5.564. The fraction of the Naturally Infected (left panel, black boxed nodes) that are Naturally Symptomatic (left panel, dashed flows) is smaller than the fraction of the vaccinated infected (right panel, black boxed nodes) that are Always Asymptomatic (right panel, dashed flows).

On the other hand, the quantity that we would use to identify this fraction under (A4) is

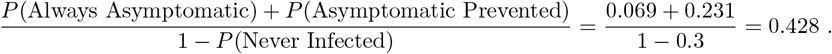

Thus, in this setting our identification formula for VE_NAI_ would be dramatically *over-estimating* the fraction of individuals who would be infected under vaccine who are Always Asymptomaticand therefore dramatically *under-estimating* the value of VE_NAI_.

The true value of the sensitivity parameter *δ* can be computed for this example as

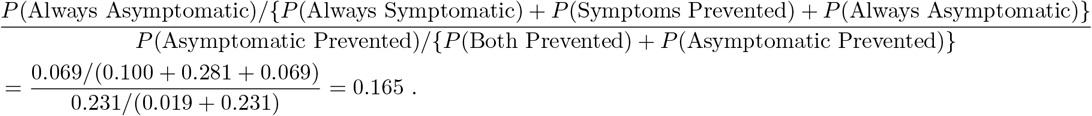

This is in line with our general observation that if the true value of *δ <* 1, then using our identification result will result in conservative inference regarding VE_NAI_.

Finally, we can consider the setting in Figure 6, with corresponding principal strata distributions shown in the right column of Table 2. In this case, we can confirm that the true fraction of Always Asymptomatic among those who would be infected under vaccine is

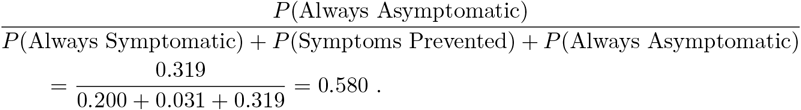

On the other hand, the quantity that we would use to identify this fraction under (A4) is

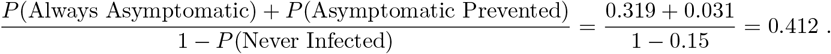

Thus, in this setting our identification formula for VE_NAI_ would be *under-estimating* the fraction of individuals who would be infected under vaccine who are Always Asymptomatic and therefore *over-estimating* the value of VE_NAI_.

The true value of the sensitivity parameter *δ* can be computed for this example as

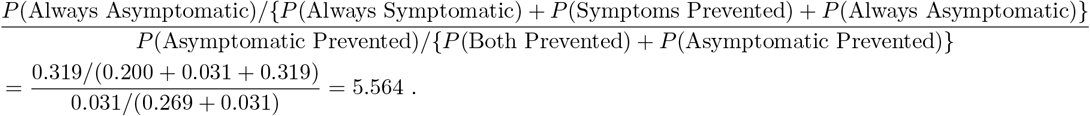

## eAppendix B. Relating VE_NAI_ to VE_Inf_

Suppose that assumption (A4) holds marginally, i.e., that *Y*_Sym_(0) ⊥ *Y*_Inf_(1) | *Y*_Inf_(0) = 1. If so, then *P* (*Y*_Sym_(0) = 0 |*Y*_Inf_(1) = 1, *Y*_Inf_(0) = 1) = *P* (*Y*_Sym_(0) = 0| *Y*_Inf_(0) = 1), which can be expressed in terms of principal strata as

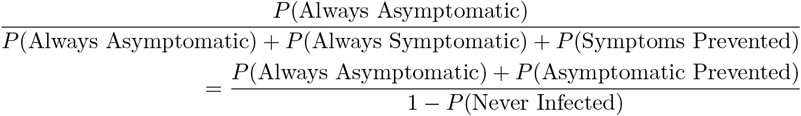

Note the left-hand side can be written as

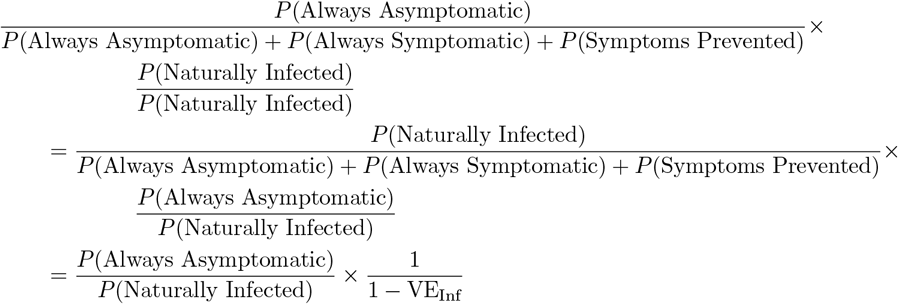

Note the right-hand side can be written as

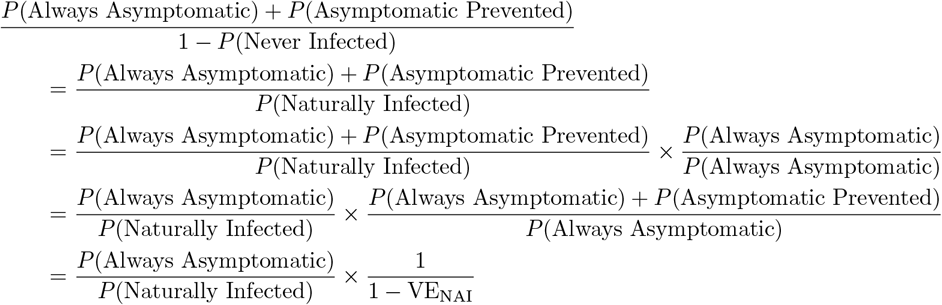

Thus, if (A4) holds marginally then VE_NAI_ = VE_Inf_.

Identical arguments can be used to show that (A4) implies that conditional VE_Inf_(*x*) = 1 − *P* (*Y*_Inf_(1) = 1 | *X* = *x*)*/P* (*Y*_Inf_(0) = 1 | *X* = *x*) is equal to conditional VE_NAI_(*x*) = 1 − *P* (*Y*_Inf_(1) = 1, *Y*_Sym_(1) = 0 | *Y*_Inf_(0) = 1, *Y*_Sym_(0) = 0, *X* = *x*). Equivalently, this means that within covariate strata, VE_Inf_ is assumed to be the same for the Naturally Asymptomatic and the Naturally Symptomatic (Figure 7). This result gives additional intuition into (A4), namely that *X* must be a sufficiently rich set of covariates such that within strata of *X*, the ability of the vaccine to prevent infection is not systematically different between those who would naturally develop symptoms and those who would not.

**Figure 7:**
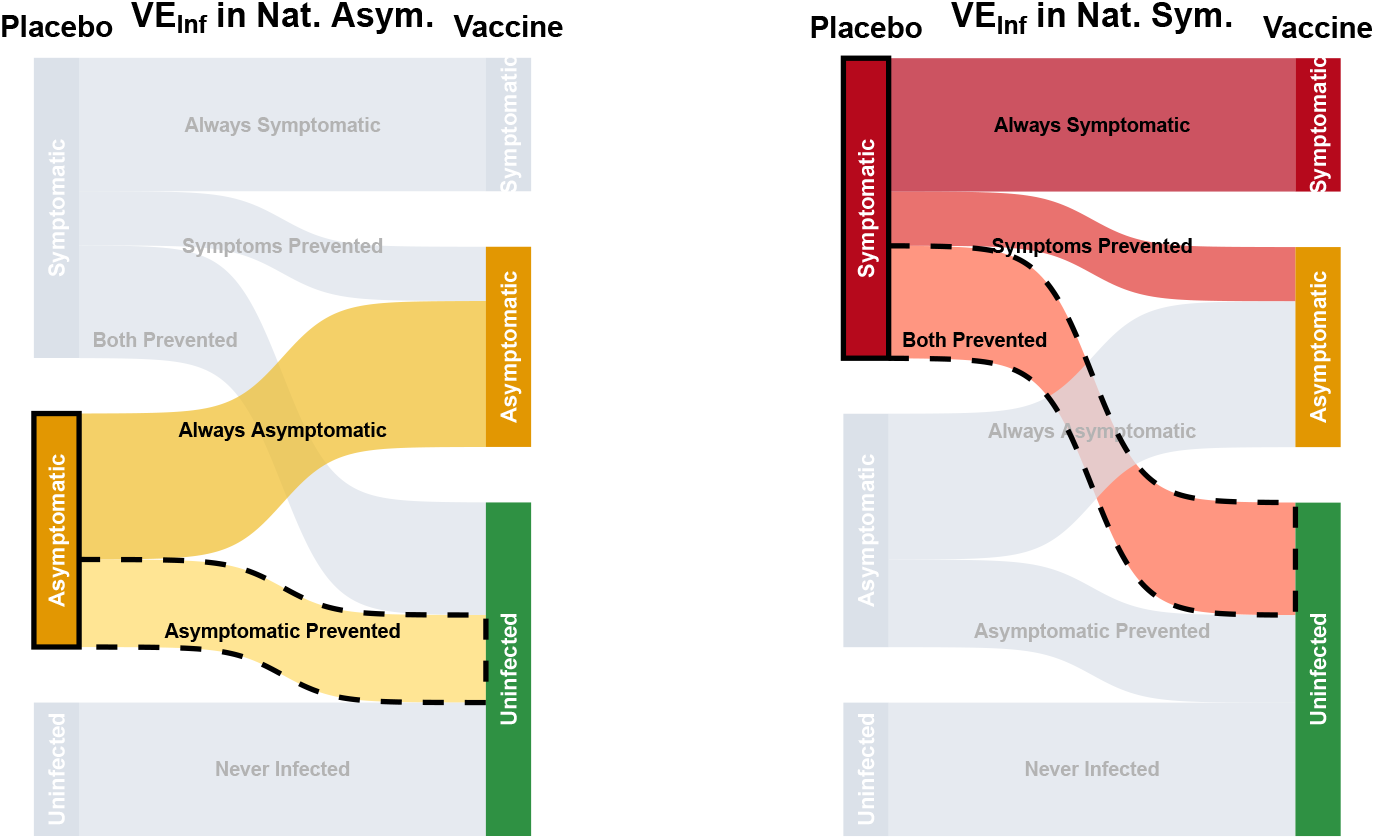
Alternative visual depiction of assumption (A4). Within strata of *X*, the fraction of the Naturally Asymptomatic (left panel, black boxed node) that have their infection prevented by vaccine (left panel, dashed flows) is equal to the fraction of the Naturally Symptomatic (right panel, black boxed node) that have their infection prevented by vaccine (right panel, dashed flows), i.e, VE_Inf_|Naturally Asymptomatic = VE_Inf_|Naturally Symptomatic.

This insight can be further reflected in an alternative formulation of the identification result, in which under (A4), we can show that VE_NAI_ is a weighted average of stratum-specific VE_Inf_, where both the weights and VE_Inf_ are easily identified in the context of a randomized trial.

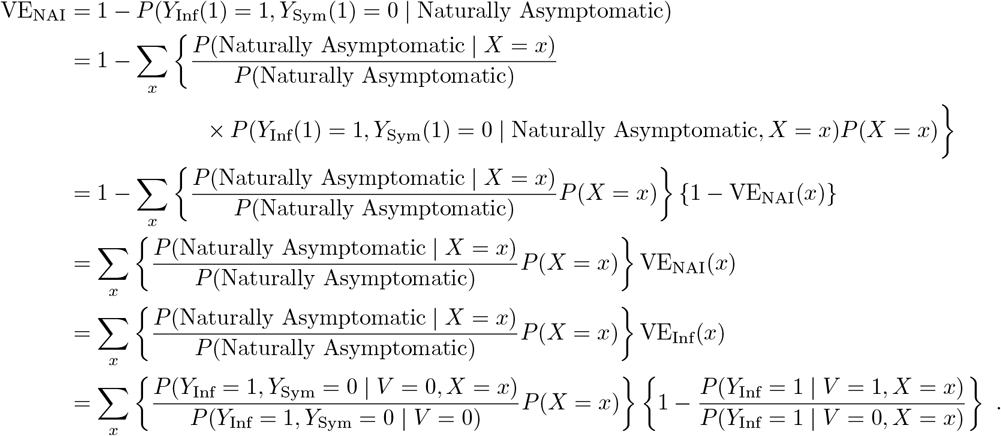

Here, the first equality is the definition of VE_NAI_. The second equality is the law of total expectation and the third is the definition of *X*-specific VE_NAI_. The fourth equality is the result of rules of joint probability implying that the weights sum to 1 over all values of *x*. The fifth equality results from an application of (A4), which we showed above implies that VE_NAI_(*x*) = VE_Inf_(*x*). The final equality results from (A0.0), (A1), and (A3). This approach to identification highlights that VE_NAI_ can be written as a weighted average of stratum-specific VE_NAI_, with weights proportional to how likely each covariate strata is to be observed among the Naturally Infected. These weights are easily identified using the observed covariate distribution and incidence rate of asymptomatic infections in the placebo arm. However, (A4) is necessary in order to identify VE_NAI_(*x*). This identification is achieved by assuming that *X* is sufficiently rich such that stratum-specific VE_NAI_(*x*) is equivalent to VE_Inf_(*x*), the latter of which is identified based on infection rates in observed covariate strata.

The fact that under (A4), VE_NAI_ can be represented as a weighted combination of VE_Inf_ implies that in the absence of moderately strong effect modification of VE_Inf_ by *X*, VE_NAI_ will often be close in magnitude to VE_Inf_. On the other hand, effect modification by *X* can lead to considerable divergence between VE_NAI_ and VE_Inf_. For example, consider the example for a hypothetical vaccine and a binary covariate *X* given in Table 3. In this example, the vaccine has a strong protective effect for the subpopulation with *X* = 0, but no effect in the subpopulation with *X* = 1. The single binary covariate *X* satisfies assumption (A4). This can be verified by confirming that within each stratum of *X*,

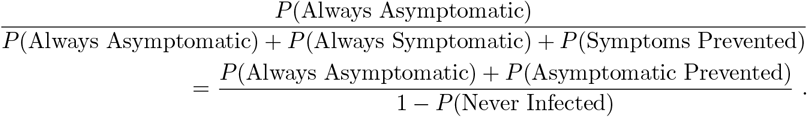

**Table 3:** Hypothetical vaccine effects and distribution of principal strata with strong effect modification by binary covariate *X*.

| | $X = 0$ | $X = 1$ |
| --- | --- | --- |
| $P(\text{Never Infected})$ | 0.900 | 0.700 |
| $P(\text{Always Symptomatic})$ | 0.005 | 0.200 |
| $P(\text{Symptoms Prevented})$ | 0.004 | 0.000 |
| $P(\text{Always Asymptomatic})$ | 0.001 | 0.100 |
| $P(\text{Both Prevented})$ | 0.081 | 0.000 |
| $P(\text{Asymptomatic Prevented})$ | 0.009 | 0.000 |
| $\text{VE}_{\text{Inf}}$ | 0.900 | 0.000 |
| $\text{VE}_{\text{Sym}}$ | 0.944 | 0.000 |
| $\text{VE}_{\text{AI}}$ | 0.500 | 0.000 |
| $\text{VE}_{\text{NAI}}$ | 0.900 | 0.000 |

For *X* = 0, both sides of the equation evaluate to 0.1, for *X* = 1 both sides evaluate to 0.333. We can also verify that within each strata of *X*, VE_NAI_ equals to VE_Inf_. However, marginally VE_NAI_ and VE_Inf_ can diverge considerably depending on the distribution of *X* (Table 4).

**Table 4:** Comparison of VE parameters in a setting with effect modification by *X* (as in Table 3), as a function of the size of the stratum in whom the vaccine has no protective effect (*X* = 1).

| $P(X = 1)$ | $\text{VE}_{\text{Sym}}$ | $\text{VE}_{\text{AI}}$ | $\text{VE}_{\text{Inf}}$ | $\text{VE}_{\text{NAI}}$ |
| --- | --- | --- | --- | --- |
| 0.01 | 92.3 | 45.4 | 87.4 | 81.7 |
| 0.10 | 75.7 | 23.7 | 67.5 | 42.6 |
| 0.25 | 54.2 | 11.5 | 45.0 | 20.8 |
| 0.50 | 29.3 | 4.5 | 22.5 | 8.2 |

These results provide insight as to why in our analysis of COVE, the estimated VE_NAI_ was very similar to the estimate of VE_Inf_ – there was little evidence of effect heterogeneity of the vaccine by any strata of covariates considered in the original trial analysis.^9^ Nevertheless, our hypothetical example above shows that similarity between VE_NAI_ and VE_Inf_ need not always be true in practice.

We can also compare VE_NAI_ to VE_Inf_ under the setting of the sensitivity analysis, where we have argued that it may be reasonable to assume *δ <* 1. This implies

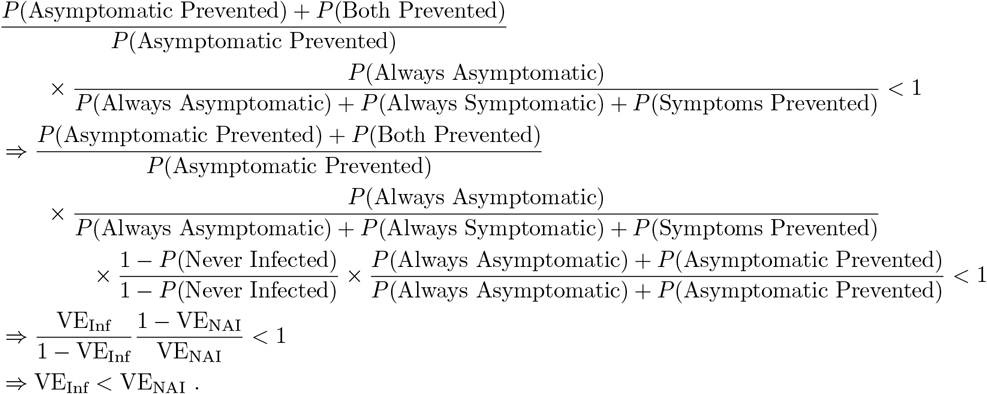

Intuitively, this corresponds to a scenario in which the vaccine’s ability to prevent infections is better among the Naturally Asymptomatic than the Naturally Symptomatic, which may be expected if the Naturally Asymptomatic have stronger immune responses and therefore are also expected to respond better to vaccine. It therefore may be most intuitive to select plausible values of *δ* based on this formulation: *δ* = VE_Inf_*/*VE_NAI_.

Furthermore, this suggests that if it is immunologically plausible to assume *δ <* 1, then VE_Inf_ could potentially be used as a lower bound for VE_NAI_ instead of VE_AI_. In settings where the Symptoms Prevented stratum is relatively large, this could tighten bounds on VE_NAI_ considerably.

## eAppendix C. Estimation with right-censored data

If individuals may be right-censored prior to the end of followup, the cumulative risk of any infection regardless of symptoms can be estimated using appropriate survival analysis methods. In our analyses, we used cause-specific Cox proportional hazards regression models to separately estimate the conditional hazard of asymptomatic and symptomatic infections. This was motivated by the fact that in many vaccine studies symptomatic infections are ascertained continuously in time (symptomatic individuals can schedule study visits at any time during follow up), while asymptomatic infections are generally ascertained at only fixed time points during follow up (e.g., through serum sampling at routinely scheduled clinic visits).

Let 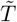 denote time to infection and Δ ∈ {0, 1, 2} denote the event type: Δ = 0: right-censored with no evidence of any infection; Δ = 1: symptomatic infection; Δ = 2: asymptomatic infection. Using these data, we fit two cause-specific Cox proportional hazards models,

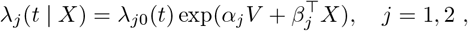

where *X* is a vector of covariates, and *j* = 1 and *j* = 2 correspond to symptomatic and asymptomatic infection, respectively. Let 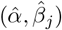 denote the estimated coefficients, and let 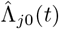 denote the Breslow estimator of the baseline cumulative hazard,

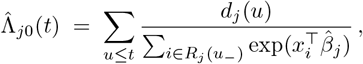

where *d*_*j*_(*u*) is the number of type-*j* events observed at time *u*, and *R*_*j*_(*u*_−_) is the corresponding risk set just prior to time *u* (i.e., individuals uncensored and event-free up to *u*). The estimated cumulative cause-specific hazard for vaccine *V* and covariates *X* at a user-selected time *t* is

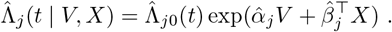

The estimated overall cumulative hazard is

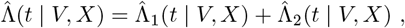

and the estimated cumulative incidence (i.e., cumulative risk) for any infection prior to time *t* is

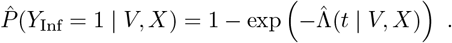

## eAppendix D. Analysis of VE against mild disease

The PROVIDE study (NCT01375647) was a randomized placebo-controlled trial of monovalent oral rotavirus vaccine (RV1) conducted in Dhaka, Bangladesh from 2011-2014.^19^ Seven hundred healthy infants were randomized 1:1 to receive two doses of RV1 or placebo at a delayed dosing schedule (10 and 17 weeks). Active twice-weekly surveillance identified diarrhea episodes among enrolled children, and rotavirus diarrhea was identified by positive stool antigen enzyme immunoassay. Diarrhea episodes with a Vesikari severity score *≥* 11 were classified as severe, while episodes ≤ 11 were classified as non-severe. We estimated VE against mild rotavirus diarrhea using generalized versions of VE_NAI_ and VE_AI_. Our analysis mirrors the primary analysis which included any episodes of rotavirus diarrhea from birth to one year of age in the per protocol population.^19^

Among 563 per-protocol participants, there were 47 and 101 episodes of rotavirus diarrhea among the vaccine and placebo recipients, respectively. 25% of episodes were severe. We characterized VE against non-severe diarrhea by estimating the analogous estimands for VE_NAI_ and VE_AI_ where *Y*_Inf_ referred to non-severe diarrhea and *Y*_Sym_ referred to severe diarrhea. In this application, *V E*_*NAI*_ estimates the effect of the vaccine to prevent non-severe diarrhea among children who would naturally have non-severe diarrhea, while VE_AI_ includes children for whom the vaccine converts disease from severe to non-severe. As expected, VE_NAI_ was larger than VE_AI_ (Table 5). Using the nonparametric Aalen-Johansen estimator, VE_NAI_ was estimated to be bounded between 42% and 71%. In a sensitivity analysis, in which *δ* ranged from ^1^ to 1.5, VE_NAI_ ranged from 60%-42% (Figure 8).

**Table 5:** Efficacy estimates and bounds for VE against mild and severe diarrhea in the PROVIDE trial at 1 year of follow-up.

| Estimate | Point Estimate | 95% CI Lower | 95% CI Upper |
| --- | --- | --- | --- |
| $VE_{\text{Inf}}$ | 0.51 | 0.33 | 0.63 |
| $VE_{\text{Sym}}$ | 0.70 | 0.36 | 0.91 |
| $VE_{\text{AI}}$ | 0.43 | 0.19 | 0.61 |
| $VE_{\text{NAI}}$ | 0.50 | 0.32 | 0.63 |
| Lower bound of $VE_{\text{NAI}}$ | 0.42 | 0.20 | 0.59 |
| Upper bound of $VE_{\text{NAI}}$ | 0.71 | 0.47 | 0.92 |

**Figure 8:**
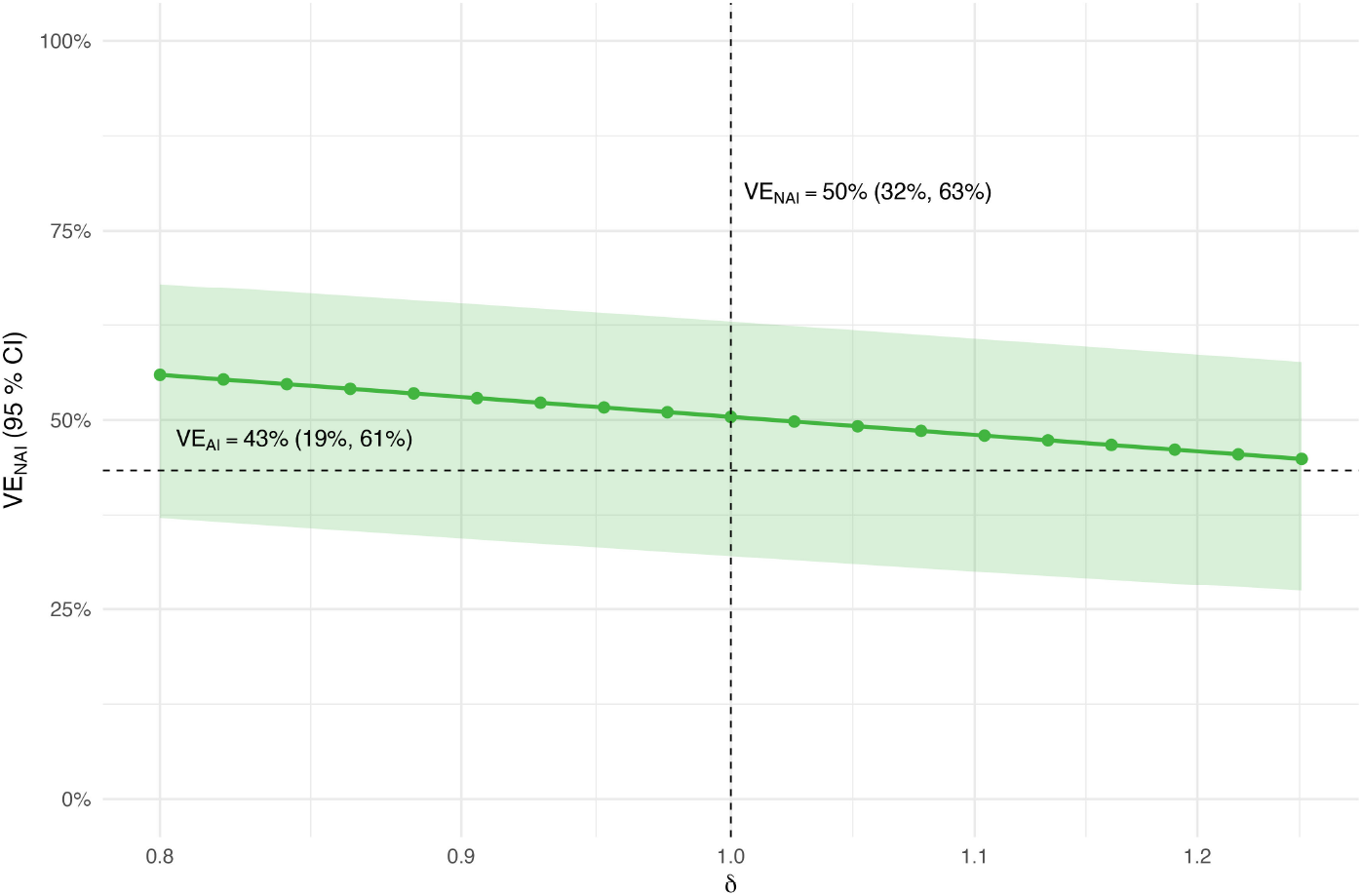
Sensitivity analysis of VE_NAI_ across *δ* ranging from 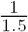 to 1.5 for PROVIDE. The horizontal dashed line indicates the estimate of VE_AI_, and the vertical dashed line indicates the estimate of VE_NAI_ at *δ* = 1.

## eAppendix E. Vaccine efficacy against progression

Under our set of assumptions, we can identify and estimate a causal version of the commonly estimated vaccine efficacy against progression:

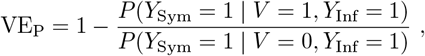

which compares rates of progression to symptoms following infection in the vaccine and placebo groups.^2^ As has been described in the literature,^26^ this estimand is clearly problematic from a causal perspective, as the infected vaccine recipients and infected placebo recipients are not comparable. This recognition has prompted the development of an alternative estimand,^26^

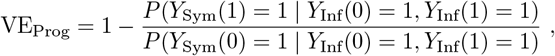

that compares the probability of progressing to symptomatic disease in the principal strata of individuals who would be infected irrespective of vaccine status. In terms of principal strata, this is

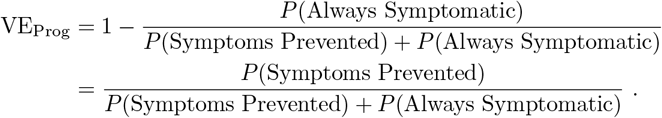

See Figure 9 for illustration. We note that this estimand – as with VE_Sym_, VE_Inf_, and VE_NAI_ – has an interpretation as the fraction of a union of principal strata that have outcomes improved by the vaccine. However, in contrast to the other estimands, the union of principal strata involved in the denominator of VE_Prog_ is not defined by the naturally occurring outcome.

**Figure 9:**
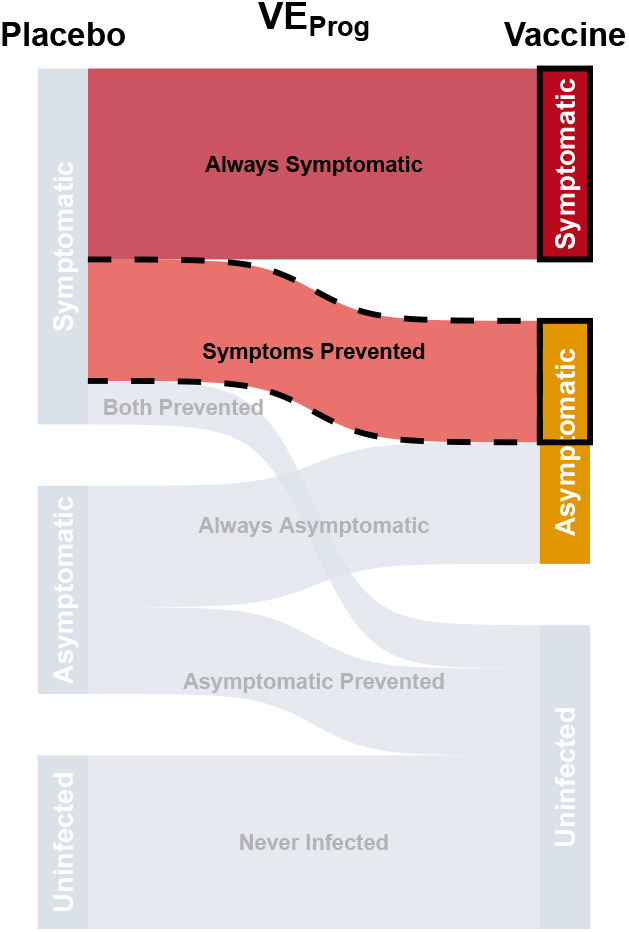
Illustration of principal strata involved in calculation of VE_Prog_.

Nevertheless, VE_Prog_ can also be identified under Assumption (A4). First, we note that *P* (*Y*_Sym_(1) = 1 | 1, *Y*_Inf_(0) = 1, *Y*_Inf_(1) = 1) is trivially identified by *P* (*Y*_Sym_ = 1 | *V* = 1, *Y*_Inf_ = 1) = ∑ _*x*_ *P* (*Y*_Sym_ = 1 | *V* = 1,*Y*_Inf_ = 1, *X* = *x*)*P* (*X* = *x* | *V* = 1, *Y*_Inf_ = 1). Next, we can write

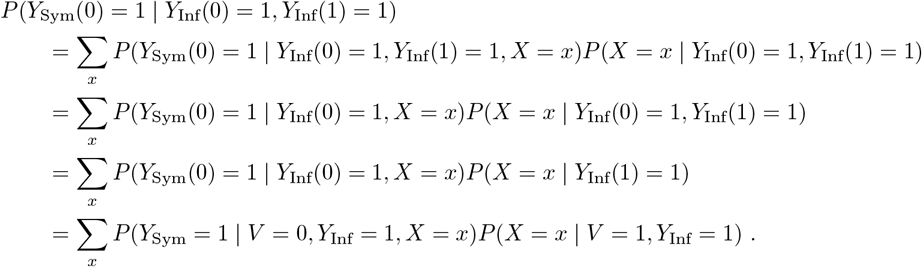

The first line follows from the law of total expectation, the second from (A4). The third line follows from (A0.2) and the final line follows from (A0.0) and (A2). Thus, we have that under the same set of assumptions needed to identify VE_NAI_, we can identify VE_Prog_ as

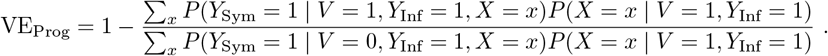

